# A Clinical Prediction Model for Sudden Cardiac Arrest Presenting as Pulseless Electrical Activity

**DOI:** 10.64898/2026.04.30.26352187

**Authors:** Harpriya Chugh, Kyndaron Reinier, Audrey Uy-Evanado, Kotoka Nakamura, Ali A. Sovari, Angelo Salvucci, Jonathan Jui, Sumeet S. Chugh

## Abstract

**Background:** The incidence of sudden cardiac arrest (SCA) manifesting as pulseless electrical activity (PEA) has increased, and survival remains extremely low. Methods for early identification and management of high-risk individuals are needed, but no clinical risk scores currently exist to predict PEA-SCA. Our objective was to develop and validate a clinical prediction model for PEA-SCA.

**Methods:** From an ongoing prospective, population-based study of SCA in Portland, Oregon (catchment pop. ≈1 M, 2002-2020), we identified PEA-SCA adults. Lifetime clinical records were compared with those of a control group with >50% prevalence of significant coronary disease. Prediction models were constructed using backwards stepwise logistic regression in a training dataset (67%) and evaluated in a validation dataset (33%). Model discrimination was assessed using receiver operating characteristic curves (C statistic). External validation was performed in a geographically distinct population in Ventura County, California (population ≈850,000, 2015-2022).

**Results:** The final clinical algorithm (PEA-Risk) incorporating 12 clinical, electrocardiogram and medication variables demonstrated strong discrimination in the training dataset (C statistic = 0.860 [95% CI: 0.838-0.881]) and remained robust in internal (C statistic = 0.832 [95% CI: 0.800-0.865]) and external validation datasets (C statistic = 0.704 [95% CI: 0.665-0.743]).

**Conclusions:** We developed and externally validated a clinical algorithm for predicting PEA-SCA. Given the low rates of successful resuscitation after PEA arrest, this risk prediction tool may enable earlier identification and prevention of PEA-SCA.

**Clinical Perspective:** *What is known:*

- The proportion of SCA presenting as pulseless electrical activity (PEA) is increasing, and survival from these events remains extremely low.
- The are no available methods for clinical risk prediction of these events.

*What the study adds:*

- The present study constructs and replicates a risk score for prediction of SCA manifesting with PEA using widely available clinical and noninvasive markers.
- These findings have implications for developing prevention and management strategies for individuals at high risk of PEA-SCA.

## INTRODUCTION

There are more than 356,000 out-of-hospital cardiac arrests annually in the U.S., nearly 90% of which are fatal, accounting for greater than 40% of cardiovascular mortality.^1^ Patients who suffer sudden cardiac arrest (SCA) can present with shockable rhythm (ventricular fibrillation / tachycardia) or non-shockable rhythm comprising pulseless electrical activity (PEA) or asystole.^2^ With improved medical and surgical therapies for ischemic heart disease and widespread use of the implantable cardioverter-defibrillator (ICD), the proportion of SCA presenting as shockable rhythm is decreasing and PEA is rising.^3–6^ Nationwide, survival from out of hospital cardiac arrests presenting with shockable rhythms is 28.7% whereas survival from a non-shockable rhythm is much lower, 6.2% ^7^. While the implantable cardioverter-defibrillator (ICD) treats SCA presenting with a shockable rhythm, the ICD is not effective for non-shockable rhythms (PEA or asystole).

Given the extremely low survival rates from PEA arrest, it is important to discover novel prevention methodologies. For example, we have recently reported a positive correlation between COPD and survival from PEA-SCA,^8^ potentially related to early treatment of hypoxia. This raises the possibility that early management of COPD exacerbations in high-risk individuals may help to prevent PEA-SCA. It would be valuable to have a clinical risk score to identify such high-risk individuals.

The Oregon SUDS (Sudden Unexpected Death Study) has prospectively ascertained SCD cases and control subjects in the general population for over two decades. The geographically distinct, but otherwise similarly designed and conducted Ventura PRESTO (Prediction of Sudden Death in Multi-ethnic Communities) study provided the opportunity to perform an external validation of such a risk score. We therefore developed and validated a clinical risk prediction score to identify patients who present with SCA manifesting as PEA.

## METHODS

### Study Design

We used a case-control design to compare SCA cases who present with PEA with control subjects who were identified and enrolled prospectively in the same period from the same geographic region.

### Study Population

#### SCA Cases

Study participants were identified (2002–2020) from the Oregon SUDS study, an ongoing investigation of SCA in the Portland, Oregon, metropolitan area. Consecutive individuals with out-of-hospital SCA were identified prospectively through collaboration with the region’s emergency medical services (EMS) system and state medical examiner’s office. A 3-physician panel adjudicated each case to determine if they met inclusion criteria for SCA of likely cardiac etiology, using all available information from patients’ existing medical records, the EMS pre-hospital report with circumstances of arrest, death certificates and autopsies when available. Patients with noncardiac causes of SCA or terminal illness were excluded (eg, trauma, overdose, pulmonary embolism, stroke, cancer not in remission, severe lung disease on home oxygen).

#### Control Subjects

Oregon SUDS control subjects were identified and enrolled prospectively in the same period from the same geographic region from multiple sources, including subjects transported by EMS for symptoms of acute coronary ischemia; patients undergoing angiography or visiting an outpatient cardiology clinic at one participating hospital; and members of a regional health maintenance organization. Control subjects were selected to have a high proportion with documented CAD but no history of ventricular arrhythmias or SCA. No matching was performed on age, sex, or other patient characteristics.

#### Inclusion criteria

All cases and controls eligible for the present analysis were ≥18 years of age with clinical history available from archived medical records. Because the focus of this analysis was to identify factors related to SCA presenting with PEA, only case subjects with an EMS-documented PEA rhythm at the time of cardiac arrest (23.4% of the total cases) were included (Figure 1). All individuals with an EMS-documented shockable rhythm, asystole or with missing rhythm were excluded (Figure 1).

**Figure 1.**
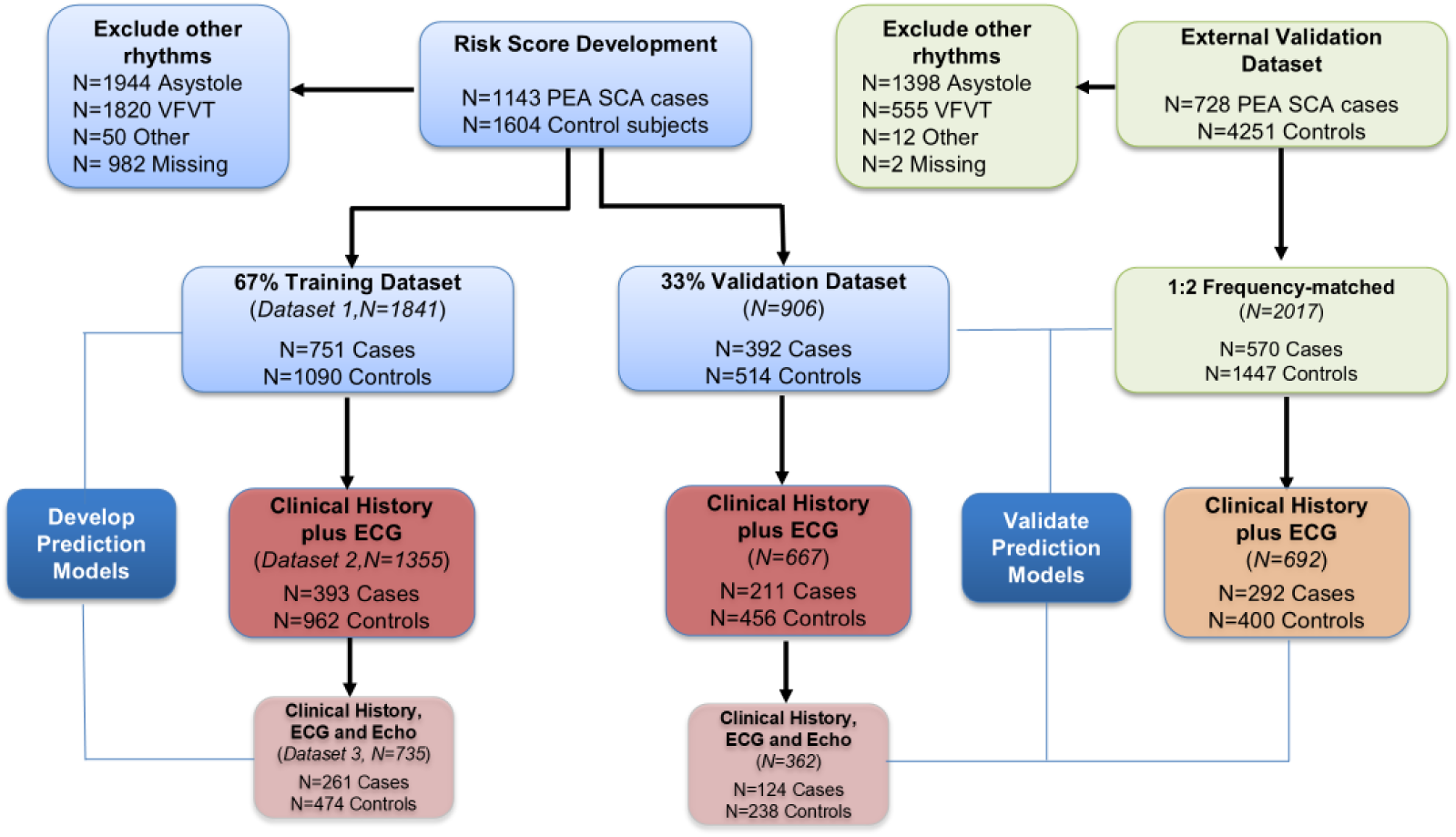
Cases and controls in the training, internal validation, and external validation datasets for prediction of SCA presenting with PEA **Legend:** The analysis was conducted among sudden cardiac arrest cases presenting with PEA. The dataset from the Oregon SUDS discovery population was divided into the training dataset (67%) for development of the prediction models, and the validation dataset (33%) to test the prediction models. Prediction models were externally validated in the geographically distinct Ventura PRESTO study. ECG = electrocardiogram; PEA = pulseless electrical activity; PRESTO = Prediction of Sudden Death in Multi-ethnic Communities; SCA = sudden cardiac arrest; SUDS = Sudden Unexpected Death Study; VF = ventricular fibrillation; VT = ventricular tachycardia.

#### External validation dataset

SCA cases for the external validation population were identified (2015–2020) from the Ventura PRESTO study, an ongoing population-based investigation of SCA in Ventura County, California. Methods for case ascertainment, adjudication, inclusion criteria, and data retrieval and definitions were identical to those used in Oregon SUDS. Validation control subjects were selected from outpatients in a health system in the region (Cedars-Sinai Medical Network, Los Angeles, California; n = 4,251). The controls were between the ages of 18 and 85. These were age and sex-matched to all PRESTO SCA cases, between the age of 18 and 85. From this larger sample, we prioritized inclusion of subjects with documented coronary disease, then selected 2 controls per PEA case using further frequency matching by race/ethnicity to the extent possible. The resulting control group had 50% documented coronary disease (Figure 1).

### Data sources

For each SCA case and control, we reviewed archived medical records to obtain a complete clinical history, including cardiovascular risk factors, cardiac tests/procedures, history of cardiac events, medications and noncardiac comorbidities. Archived 12-lead electrocardiograms (ECGs) were obtained (before and unrelated to the SCA event for cases). When more than 1 ECG was available, the one proximate to the SCA event (or the ascertainment date for controls) was selected. Archived echocardiograms were obtained in a similar manner. Available clinical history was required for inclusion in the analysis dataset. ECGs and echocardiograms were available for a subset of participants.

### Statistical methods

#### Study variables

To construct the risk score model, we considered patient demographics (age, sex, and race/ethnicity), and the following clinical predictors (n=16) and medications (n=3) based on prior association with SCA: body mass index (BMI, calculated as weight [kg]/height [m]2), diabetes mellitus (DM), hypertension, heart failure (HF), stroke, atrial fibrillation, history of myocardial infarction (MI), chronic obstructive pulmonary disease (COPD), chronic kidney disease (CKD), seizure disorder, history of syncope, anemia, sleep apnea, peripheral vascular disease (PVD), mood disorder, cancer, and medications (anti-psychotic, anti-depressants and beta blockers)^9–23^.

ECG predictors (n = 11) included heart rate, corrected QT interval (Bazett’s), Tpeak-Tend interval, delayed intrinsicoid deflection, left ventricular hypertrophy (LVH) by ECG, QRS duration, QRST angle, delayed QRS transition, left bundle branch block, prolonged PR interval, and left atrial enlargement.^24,25^

Echocardiographic predictors (n = 6) included LVEF, LVH defined as an LV mass index >134 g/m2 for men and 110 g/m2 for women, moderate to severe mitral regurgitation, moderate to severe aortic stenosis, moderate to severe aortic insufficiency and mitral valve prolapse.^26–29^

#### Training and internal validation sets

We divided the Oregon SUDS case-control data into a training dataset (67%) and a validation dataset (33%), each with 3 separate analysis data subsets: Dataset 1 (all subjects), dataset 2 (restricted to subset with ECG data, regardless of echocardiogram availability), and dataset 3 (restricted to subset with both ECG and echocardiogram data available) (Figure1). Datasets 2 and 3 were constructed using stratified random sampling of Dataset 1, based on ECG and echocardiogram data availability, pooling cases and controls to ensure that both the training and validation datasets had approximately equal proportions of ECG and echo data available for both cases and controls.

#### Preliminary case-control analysis

We used Student’s t-tests and chi-square tests to compare means and frequencies of each predictor in cases and controls in the training and validation datasets 1 through 3. SAS version 9.4 (SAS Institute) was used for all analyses.

#### Development of prediction models in the training dataset

We used backwards stepwise logistic regression in the training dataset to build prediction models, retaining variables with P < 0.20. Three separate models were constructed: model 1 (clinical variables only from dataset 1); model 2 (retained clinical variables from model 1 and ECG predictors using dataset 2); and model 3 (retained clinical variables from model 1, retained ECG variables from model 2, and echocardiogram predictors, using dataset 3). No echocardiogram variables were significant in model 3 and so we retained model 1 and model 2 for further validation.

Receiver operating characteristic (ROC) curves (C statistics) were used to evaluate model discrimination (ie, the ability of the model to separate individuals who experienced SCA from those who did not).^30^ To obtain parsimonious, easily applicable models, we changed the significance threshold to p <0.05, removed predictors with an odds ratio (OR) below 1.0 and made age a categorical variable (age ≤ 60, >60 to 70 years, >70 to 80 and >80 years), creating our final PEA-Risk models.

#### Internal validation

We applied the fixed beta coefficients from the logistic regression models developed in the training datasets to the validation datasets using the “score” statement in SAS, separately for model 1 and model 2. For each model, the C statistic was interpreted as the overall performance of the prediction model as applied to the respective validation dataset.

#### Risk score development

The risk score (PEA-Risk) was developed using variables retained in model 2 (clinical and ECG predictors). Points were assigned based on the OR for each predictor from the training dataset, rounded to the nearest tenth.

ORs associated with a 1-unit increase in the risk score, and by risk categories, were calculated in the training and validation datasets separately to evaluate the risk score’s utility. In addition, we divided the PEA-Risk score into 3 risk categories based on the case-control pooled group (low: ≤ median, medium: > median to 75^th^ percentile, and high: > 75^th^ percentile). Finally, in the combined training and validation datasets, we evaluated the consistency of risk score performance in population subgroups by testing for interaction between the PEA-Risk score and sex, age, and history of CAD and calculated ORs by PEA-Risk categories stratified by sex, age, and history of CAD. *External Validation*

We applied the fixed beta coefficients from the training logistic regression models to the external validation datasets using the “score” statement in SAS and used C statistics to evaluate the performance of the prediction model in the external validation dataset for models 1 and 2. PEA-Risk points were then assigned to each individual in the external validation dataset based on their clinical and ECG profile to calculate the OR associated with a 1-unit increase in the risk score and ORs by risk score categories.

## Results

### Demographics of subjects

A total of 1143 SCA cases (62% male, mean age 70 ± 16 years) ascertained in the Oregon SUDS from 2002 to 2020 presented with PEA and met criteria for analysis (Figure 1). A total of 1,604 control subjects (67% male, mean age 65 ± 13 years) were enrolled during the same period and met criteria for inclusion in this analysis. The full case-control dataset was divided into a training dataset (n=1841) with 751 cases and 1,090 controls, and an internal validation dataset (n=906) with 392 cases and 514 controls (Figure 1). Dataset 2 consisting of the subset with ECG available (n=1355 in the training and n=667 in the validation datasets) became our primary dataset.

### Case and control characteristics: training dataset

In dataset 2, cases were older and several cardiovascular risk factors, prevalent cardiac disease, and noncardiac comorbidities were significantly more common among cases than controls including hypertension, history of MI, DM, prevalent HF, and a history of stroke, atrial fibrillation, syncope, COPD, CKD, anemia, PVD, hypothyroid and mood disorder (Table1; p <0.02). Use of anti-psychotic and anti-depressant medications was significantly more common in cases than controls (p <0.02). All abnormal ECG findings were significantly more prevalent among cases (p <0.02) except for left atrial enlargement (Table 1). In the subset that had echocardiograms, reduced ejection fraction, LVH and mitral regurgitation were significantly more prevalent in the cases than controls (Table 1).

**Table 1:**
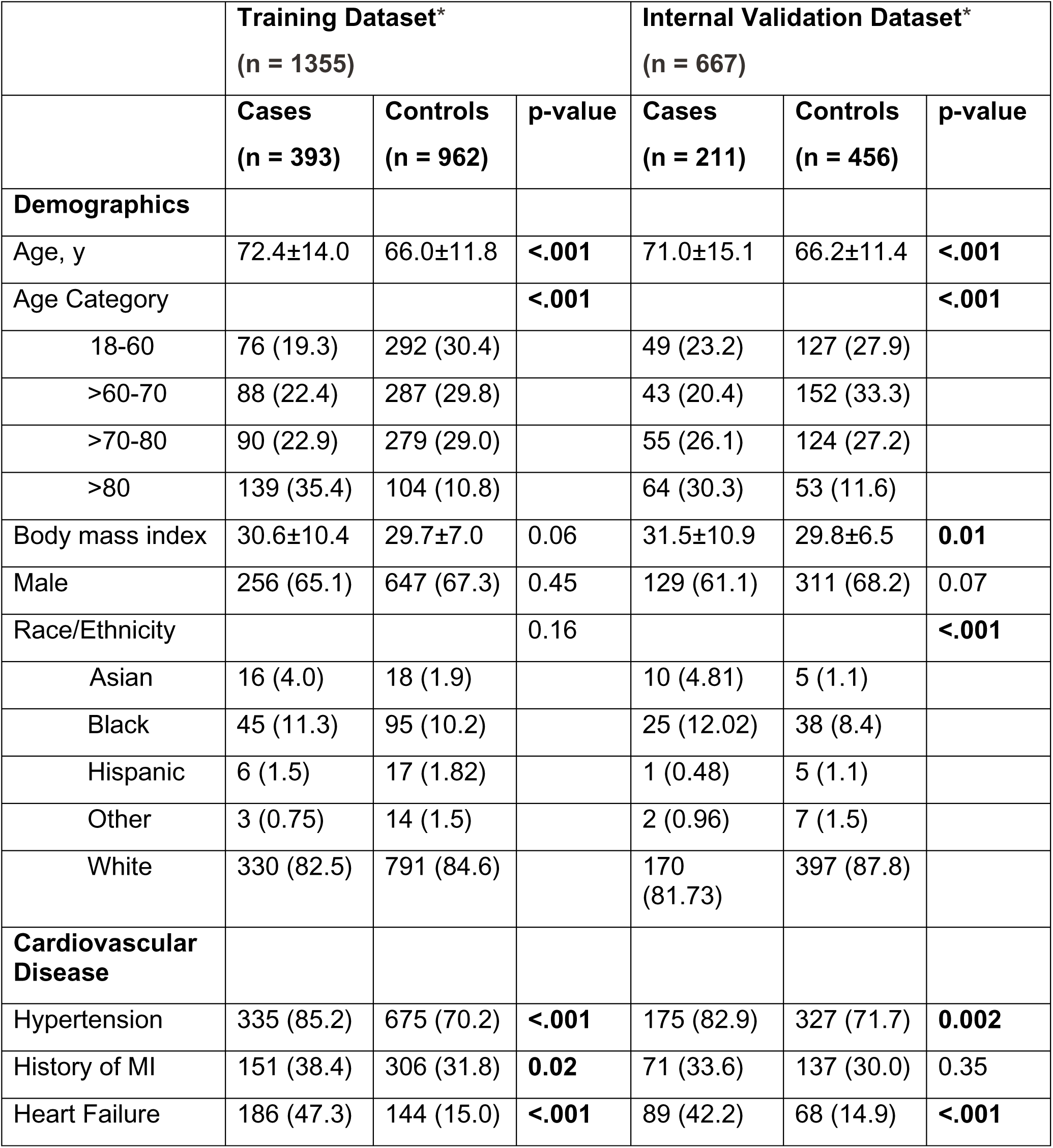
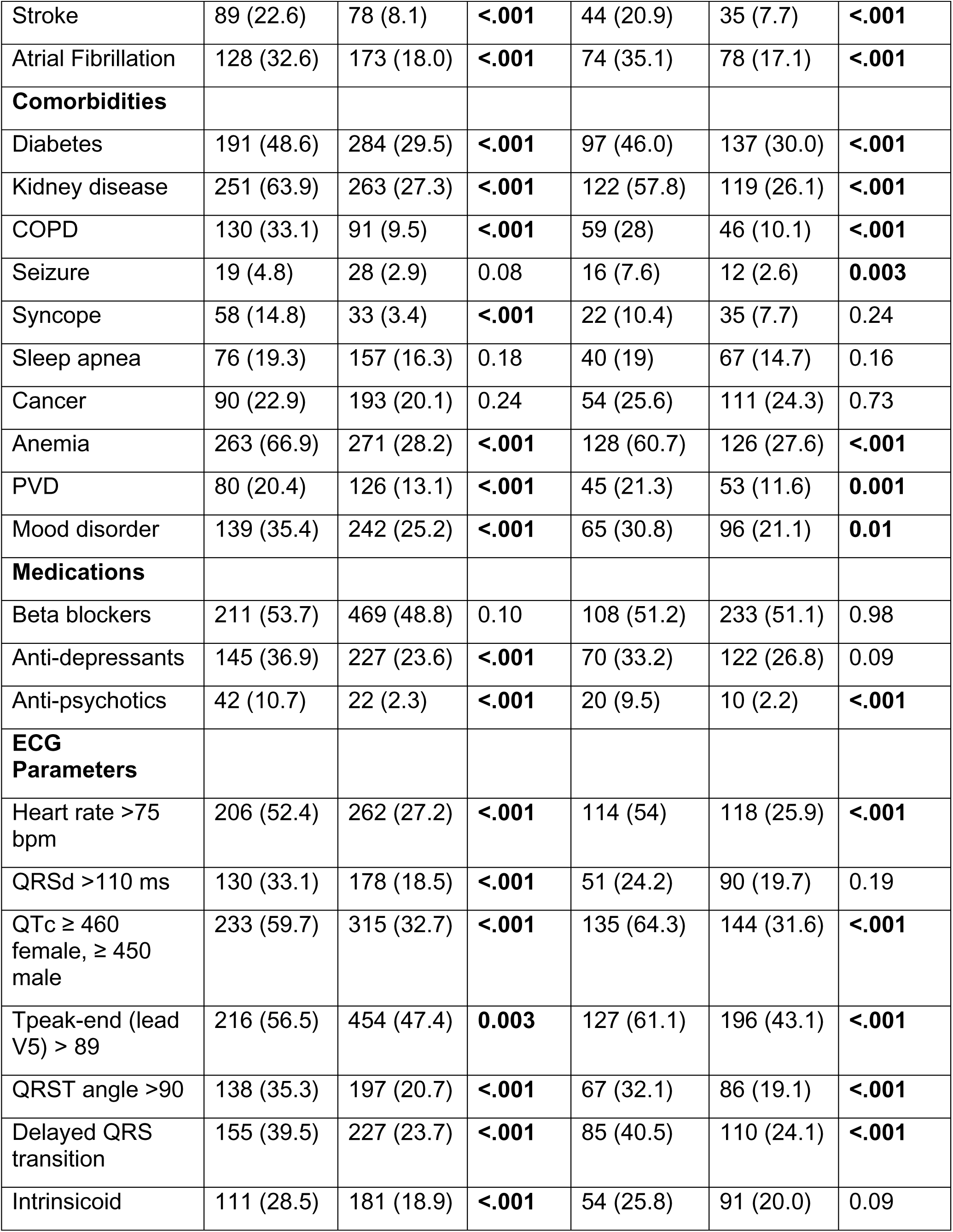
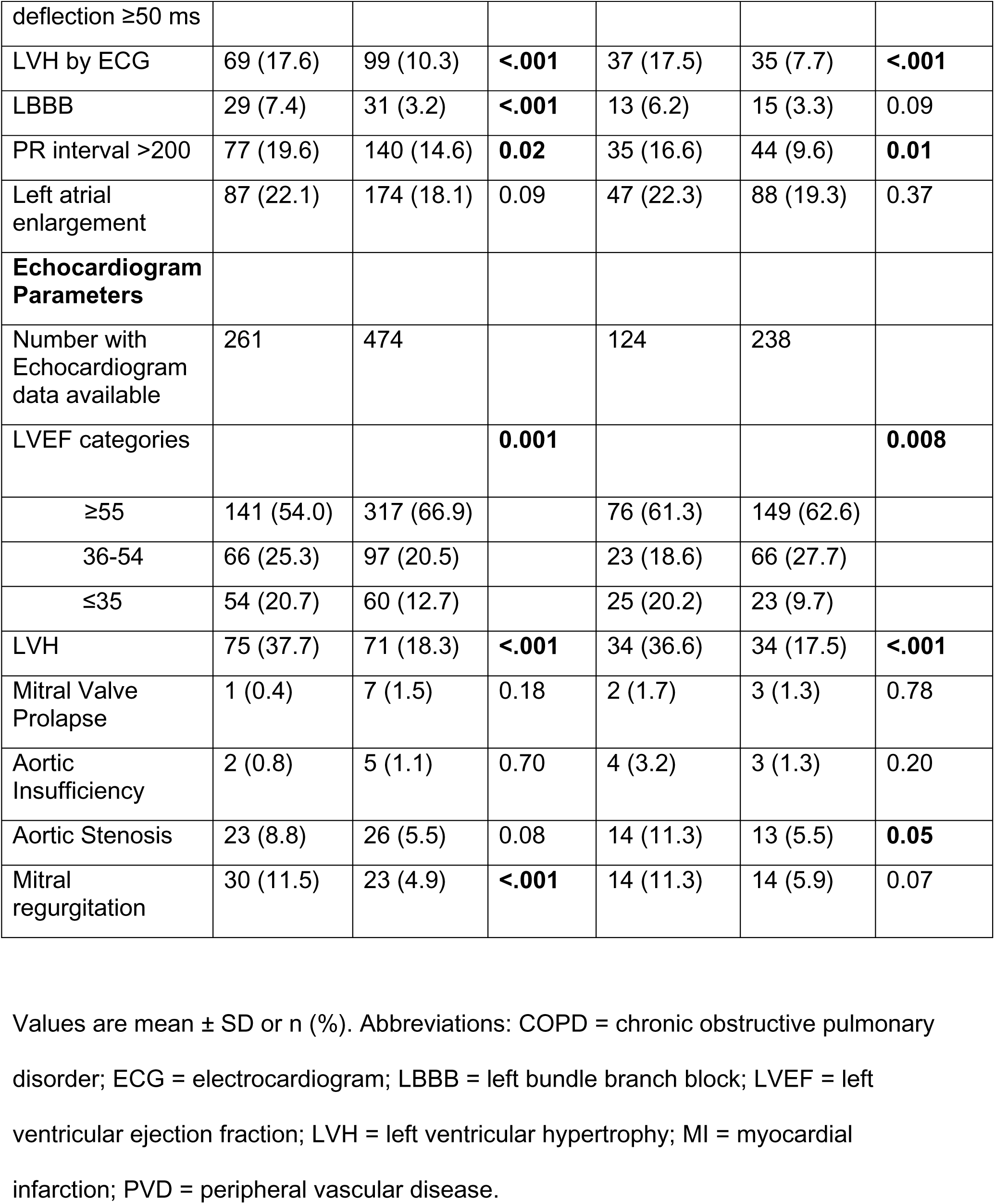
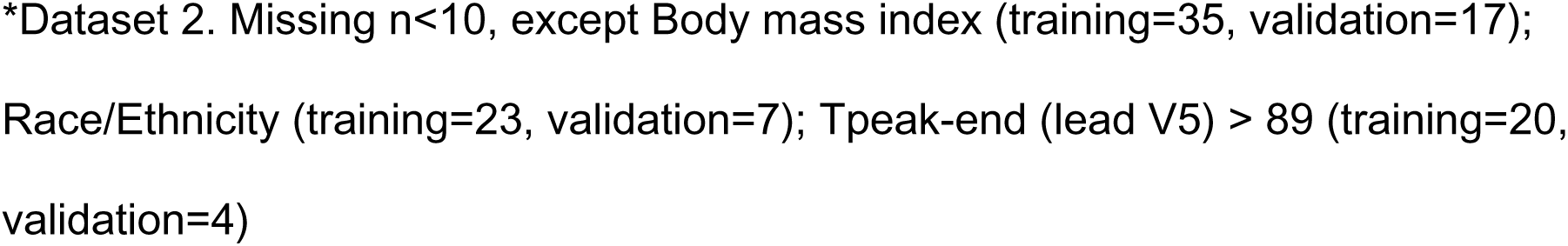
Univariate comparisons of Oregon SUDS PEA-SCA cases and controls with complete data for demographics, clinical phenotypes, and ECG variables.

### Multivariable prediction models

After applying the criteria to obtain parsimonious models (variables with p<0.05 and OR >1.0), in model 1 most of the clinical variables and medications were retained (Table 2). Sex, race, mood disorder, and seizure disorder dropped out of the model. MI, atrial fibrillation, hypertension, sleep apnea, cancer, PVD, and beta blocker use had odds ratios below 1.0. Model 2 with clinical and ECG variables had similar predictors to Model 1 with the exception that body mass index dropped out and stroke and use of anti-depressants became significant. Increased heart rate, Tpeak to Tend interval and delayed QRS transition zone were retained from the ECG parameters (Table 2). In model 3 with clinical, ECG and echocardiogram variables, none of the echocardiogram markers met criteria for retention in the model (Supplementary Table 1). Based on this finding, model 3 was not carried forward.

**Table 2:** Multivariable logistic regression PEA-Risk prediction models in the training dataset in SUDS.

|  | **Clinical**<br>**Model 1‡**<br>**(n = 1841)** |  | **Clinical + ECG**<br>**Model 2§**<br>**(n = 1355)** |  |
| --- | --- | --- | --- | --- |
| AUC | 0.746 [0.723-0.769] |  | 0.860 [0.839-0.882] |  |
|  | Adjusted OR*<br>[95% CI] | Points for<br>Risk Score | Adjusted OR†<br>[95% CI] | Points for<br>Risk Score |
| Demographics |  |  |  |  |
| Age |  |  |  |  |
| 18-60 (Reference) |  |  |  |  |
| >60-70 | 0.85 [0.64-1.12] |  | 1.05 [0.69-1.61] |  |
| >70-80 | 0.90 [0.67-1.20] |  | 1.00 [0.66-1.54] |  |
| >80 | 2.78 [2.01-3.84] | 2.8 | 3.88 [2.47-6.08] | 3.9 |
| Body Mass Index |  |  |  |  |
| <25 | 1.44 [1.10-1.89] | 1.4 |  |  |
| 25-29.9 (Reference) |  |  |  |  |
| 30-34.9 | 0.78 [0.57-1.07] |  |  |  |
| ≥35 | 1.76 [1.30-2.38] | 1.8 |  |  |
| missing | 1.55 [0.89-2.72] |  |  |  |
| Cardiovascular Disease |  |  |  |  |
| Heart Failure | 2.06 [1.58-2.69] | 2.1 | 2.21 [1.57-3.10] | 2.2 |
| Stroke |  |  | 1.73 [1.14-2.62] | 1.7 |
| Comorbidities |  |  |  |  |
| Kidney Disease | 1.38 [1.08-1.75] | 1.4 | 2.05 [1.49-2.83] | 2.1 |
| COPD | 2.36 [1.74-3.20] | 2.4 | 2.97 [2.03-4.34] | 3 |
| Syncope | 2.39 [1.52-3.76] | 2.4 | 3.21 [1.85-5.58] | 3.2 |
| Anemia | 1.42 [1.12-1.79] | 1.4 | 2.71 [1.99-3.69] | 2.7 |
| Medications |  |  |  |  |
| Anti-depressants |  |  | 1.59 [1.14-2.20] | 1.6 |
| Anti-psychotics | 4.32 [2.58-7.22] | 4.3 | 3.53 [1.81-6.91] | 3.5 |
| ECG Parameters |  |  |  |  |
| Heart rate >75 bpm |  |  | 2.84 [2.09-3.86] | 2.8 |
| Tpeak-end (lead V5) > 89 |  |  | 1.75 [1.30-2.37] | 1.8 |
| Delayed QRS transition |  |  | 1.42 [1.03-1.95] | 1.4 |

Abbreviations: OR = odds ratio; AUC= Area Under the Curve; COPD = chronic
obstructive pulmonary disorder; ECG = electrocardiogram
\* ORs are adjusted for all other clinical variables in the model.
† ORs are adjusted for all other clinical and ECG variables in the model.
‡ Model created in training dataset with data for clinical phenotypes using clinical
variables (dataset 1). The points for the risk score constitute the secondary PEA-Risk
score.
§ Model created in training dataset with data for clinical phenotypes, and ECG variables
using clinical variables and ECG variables (dataset 2). The points for the risk score
constitute the PEA-Risk score.

Model 2 (clinical + ECG) had significantly higher discrimination than model 1 (clinical variables only) (C statistic 0.860 vs 0.746, p<0.0001) (Figure 2). Model 2 was treated as the primary model. Table 2 shows the final model 1 and model 2.

**Figure 2.**
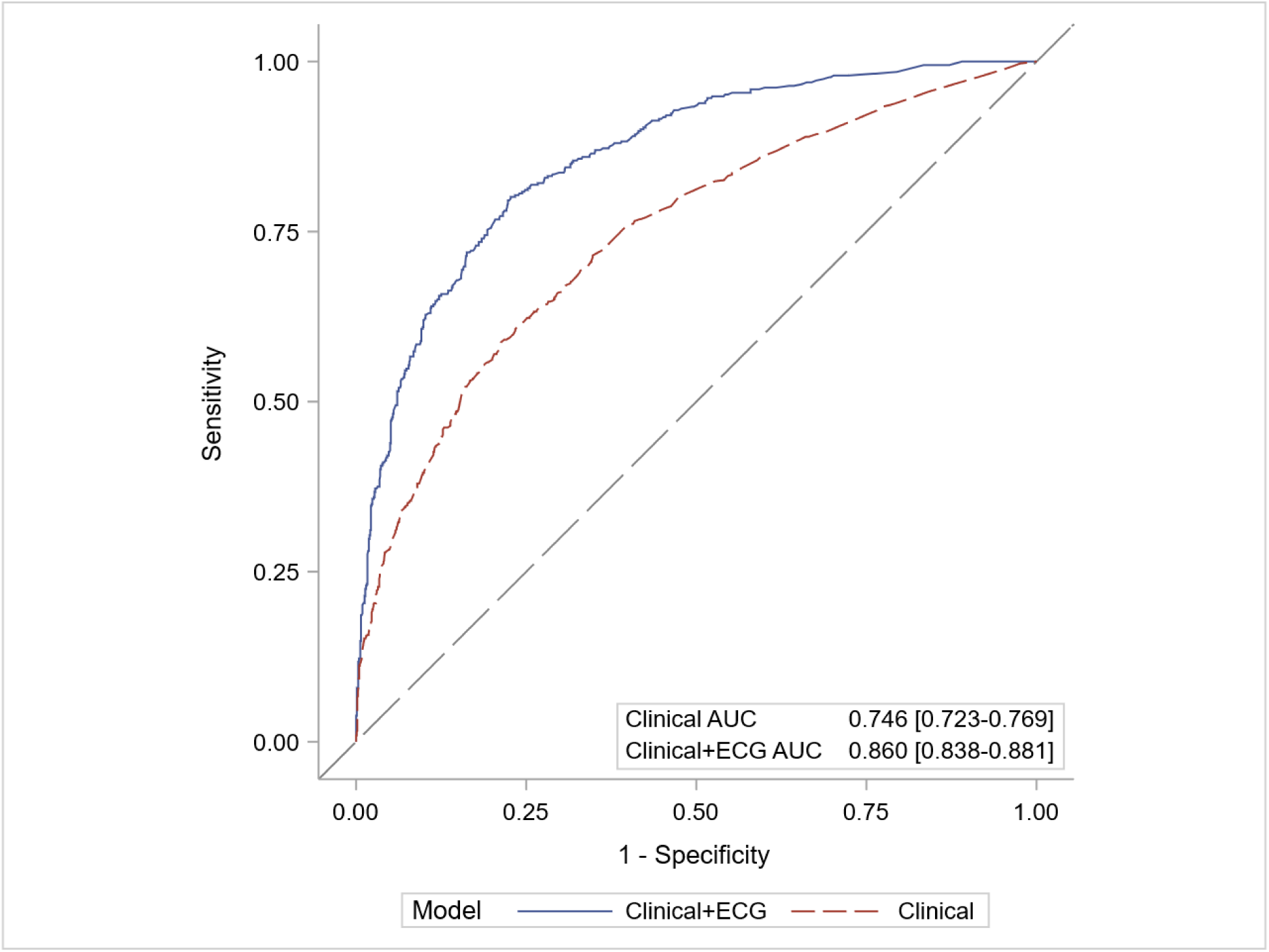
ROC Curves for models with clinical predictors and a combination of clinical and ECG predictors **Legend**: Prediction models shown include (model 1) clinical (medical history) predictors only run on 1841 subjects; (model 2) clinical plus ECG run on 1355 subjects. ECG = electrocardiogram; ROC = receiver operating curve; AUC = Area under the curve

#### Internal validation of prediction models

When the fixed coefficients from the training dataset were applied to the held-out validation dataset, the C statistics indicated good discrimination (model 1, 0.730 [0.697-0.763] and model 2, 0.832 [0.800-0.865] (Table 3).

**Table 3:** C Statistics Evaluating the Performance of PEA-Risk Prediction Models in the Training and Validation Datasets.

|  | **C Statistic (95% CI)** |  |  |
| --- | --- | --- | --- |
|  | **Training dataset** | **Internal Validation** | **External Validation** |
| Clinical variables only | (n=1841)<br>0.746 [0.723-0.769] | (n=906)<br>0.730 [0.697-0.763] | (n=2017)<br>0.680 [0.654-0.707] |
| Clinical + ECG | (n=1355)<br>0.860 [0.838-0.881] | (n=667)<br>0.832 [0.800-0.865] | (n=692)<br>0.704 [0.665-0.743] |

Abbreviations: ECG = electrocardiogram

#### Risk score (PEA-Risk)

The points used to construct the primary (model 2) and the secondary (model 1) PEA-Risk score are shown in Table 2. For model 2, a total of 12 components resulted in a median score of 6.1 (0 minimum to 24.4 maximum) in the training dataset and 6.1 (0 to 21.5) in the internal validation dataset. The median risk score among cases in the training and validation datasets was 11.4 (1.4 to 24.4) and 10.3 respectively (0 to 21.5), whereas among controls it was 4.6 (0 to 19.2) in the training dataset and 4.5 (0 to 19.7) in the validation dataset (Figure 3). A 1-unit increase in the risk score was associated with a 42% increase in odds of PEA-SCA (OR: 1.42; 95% CI: 1.37-1.47) with similar results in the internal validation dataset (OR: 1.33; 95% CI: 1.27-1.40). ORs by PEA-Risk categories, with the lowest category as reference, were also consistent in the training and validation datasets, with the highest risk score category associated with a 16-fold to 29-fold higher odds of PEA than the lowest risk category (Table 4).

**Figure 3.**
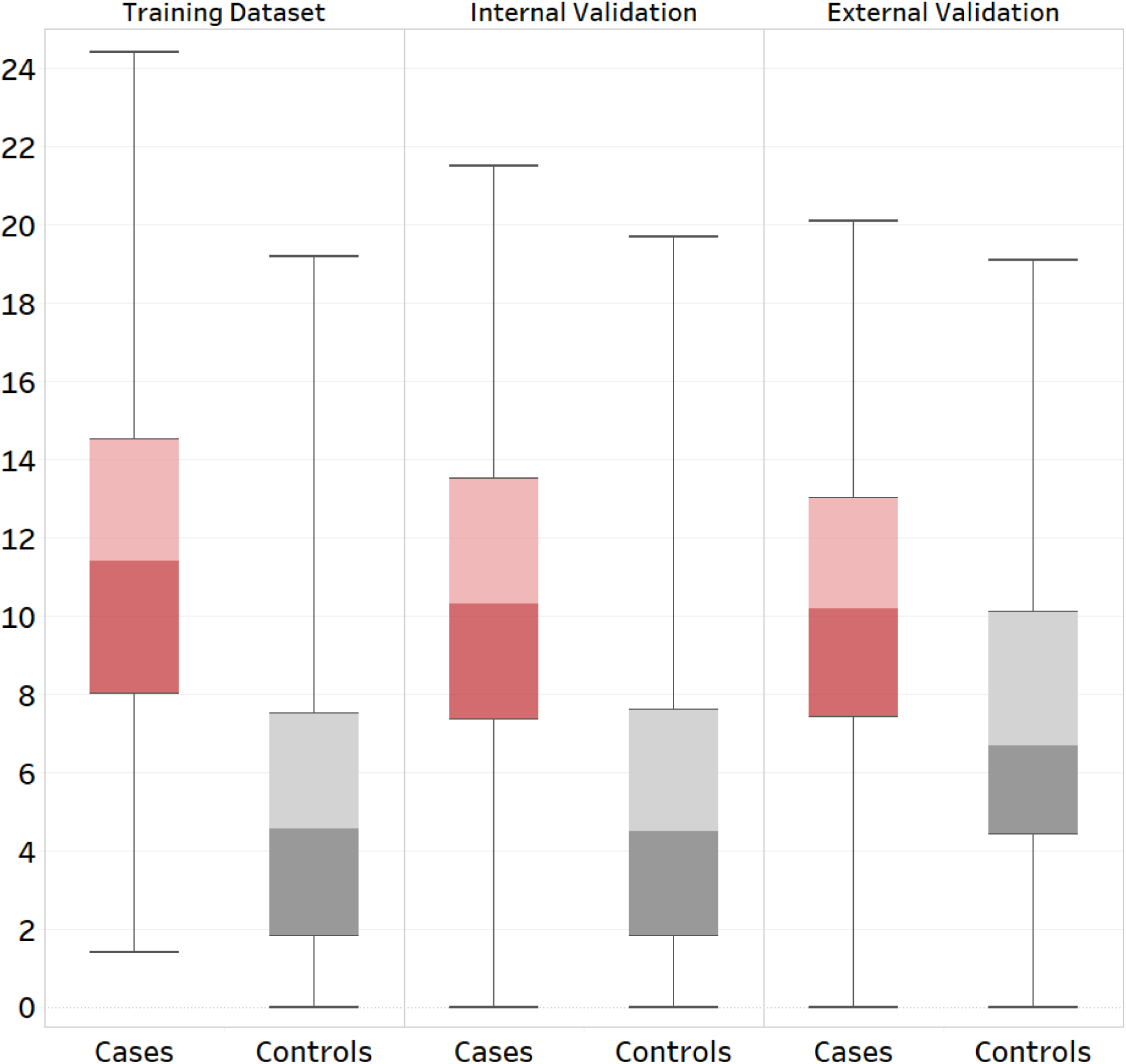
Distribution of the PEA-Risk score in cases and controls in the training and validation datasets **Legend** Box plot showing the median (line where the colors change), interquartile range (box), and minimum and maximum values (whiskers) of PEA-Risk score among cases and controls in the training (n=1355), internal validation (n=667) and external validation (n=692) datasets.

**Table 4:** Risk prediction by PEA-Risk categories in the Training and Validation Datasets*.

|  | OR [95% CI], by PEA-Risk categories |  |  |
| --- | --- | --- | --- |
|  | Training | Validation |  |
| PEA-Risk Categories | (n = 1355) | Internal<br>(n = 667) | External<br>(n = 692) |
| Low: (0.0-6.1 points)<br>(Reference) | 1.00 | 1.00 | 1.00 |
| Medium: (6.2-10.1 points) | 4.87 [3.39-6.99] | 6.40 [3.99-10.24] | 2.46 [1.62-3.71] |
| High: (10.2-24.4 points) | 29.40 [20.38-42.41] | 15.52 [9.7-24.83] | 5.19 [3.46-7.80] |

\*Dataset 2 (clinical and ECG data). Low: <median score value; Medium: >median to 75<sup>th</sup> percentile; High: >75<sup>th</sup> percentile. Abbreviations: OR = odds ratio; PEA =Pulseless Electrical Activity

#### External validation

Characteristics of the external validation population are shown in Supplementary Table 2. In the external validation dataset, the C statistic indicated good discrimination (C statistic 0.704 [0.665-0.743]) for model 2 (Table 3). Median risk scores in the external validation (10.2 among the cases and 6.7 among the controls) were similar to those in the training dataset (Figure 3). A 1-unit increase in the PEA-risk score was associated with a 20% increase in odds of PEA-SCA (OR: 1.20; 95% CI: 1.15-1.25). *Performance of PEA-Risk in population subgroups*

Based on the consistent results in the training and validation datasets, we evaluated the performance of the score by population subgroup in the combined Oregon SUDS dataset (Figure 4). Models with interaction terms between the score and age, sex and CAD revealed no significant interaction, indicating similar performance across subgroups. ORs increased across risk categories (with the lowest category as reference) for all subsets by age, sex, and CAD.

**Figure 4.**
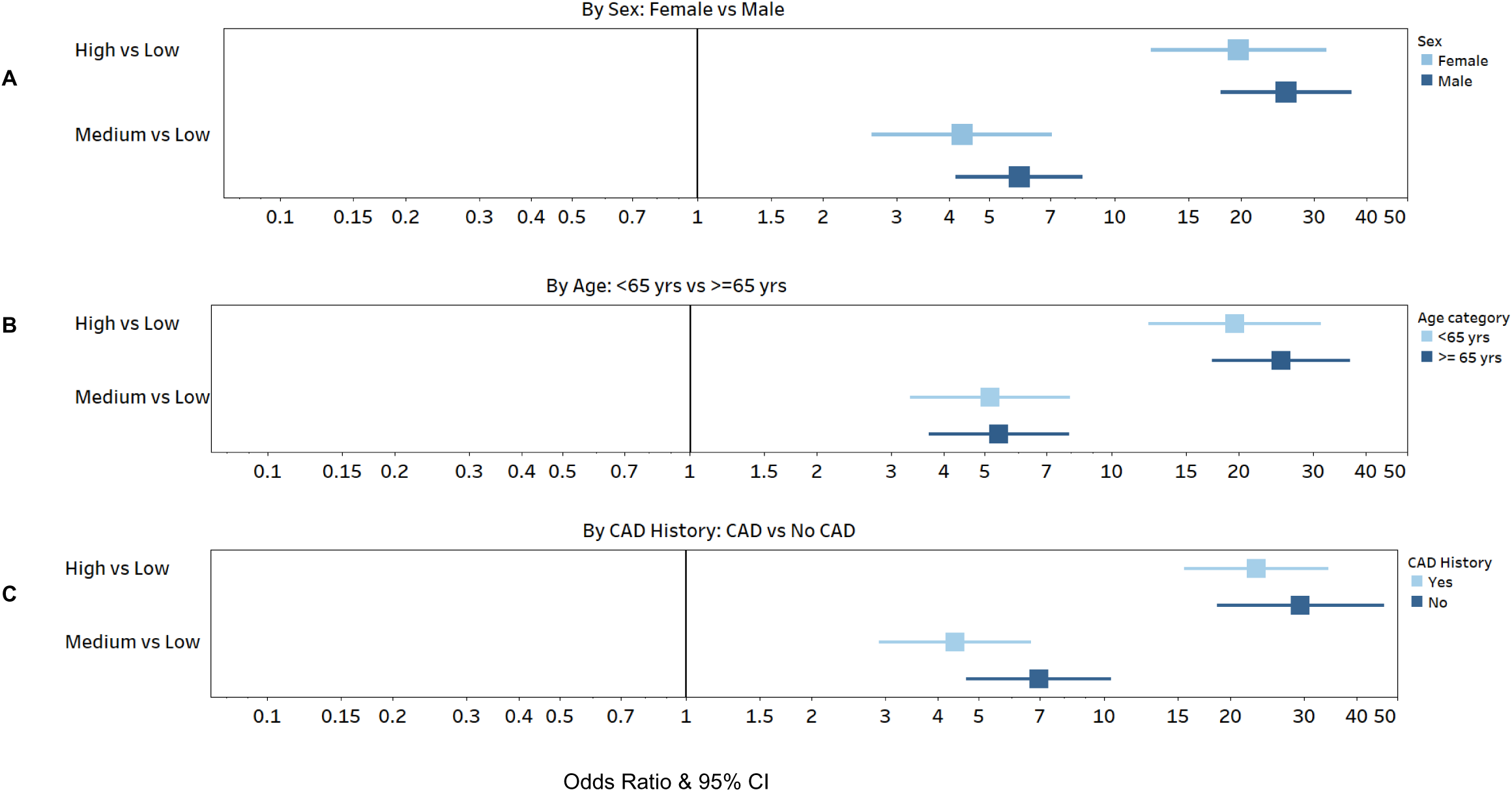
Performance of the PEA-Risk Score to predict PEA-SCA in population subgroups in the combined training and internal validation dataset **Legend** Odds Ratio and 95%CIs by PEA-Risk Categories by sex (A), by age (B) and history of coronary artery disease (CAD) (C), in the combined dataset (n=2022, dataset 2 with clinical and ECG variables). PEA-Risk categories were based on the PEA-Risk score in the case-control pooled group (Low (Reference): 0-6.1; Medium: 6.2-10.1 and High:10.2-24.4).

## DISCUSSION

We report an algorithm for prediction of out-of-hospital SCA manifesting with EMS documented PEA (PEA-Risk), constructed with 12 clinical variables, medications and ECG variables. PEA-Risk displayed good discrimination in the training dataset as well as the internal validation dataset. External validation was successfully conducted in a geographically distinct population. PEA-risk performed equally well across subpopulations stratified by sex, age and coronary artery disease. Based on the odds ratio, the key components of PEA-risk score were age >80, history of COPD, syncope and use of anti-psychotic medications. These were followed by heart failure, kidney disease, anemia and heart rate >75 bpm. Stroke, use of anti-depressants, Tpeak to Tend >89, and delayed QRS transition on the ECG also contributed to the model.

A secondary risk prediction model comprised of only clinical and medication variables performed moderately well.

To our knowledge PEA-risk is the first clinical algorithm assembled for the prediction of SCA presenting with documented PEA. We have previously evaluated differences between VF and PEA manifestations of SCA.^10,11,31^ We recently discovered and validated a clinical algorithm that identifies patients who present with SCA manifesting as VF/VT (VFRisk).^9,32^ Comparing the 2 risk scores, older age, CKD, anemia, use of anti-depressant and anti-psychotic medications, and presence of delayed QRS transition zone on the ECG were included in PEA-Risk, but not present in the VFRisk score. COPD, syncope and increased heart rate are present in both PEA-Risk and VFRisk with higher weight in PEA-Risk as compared to VFRisk. Heart failure, stroke and Tpeak to Tend on the ECG had similar weight in both the scores. History of myocardial infarction, atrial fibrillation, diabetes, seizure, prolonged QTc and delayed intrinsicoid deflection on the ECG, and LVH were present in the VFRisk but not in the PEA-Risk score.

Published studies support the inclusion of individual variables within PEA-Risk.

The association between COPD and increased risk of SCD has been reported consistently. ^9,17,33,34^ In our previous study we found that COPD is more commonly found in SCA presenting with PEA than VF/VT.^31^ In OHCA primarily in a COPD setting, the initial rhythm was PEA in 35% and VT/VF in 8%.^35^ The association between older age and PEA has also been reported.^31,36^ Similar to our current study, Kauppila et.al found that antipsychotics (OR 4.27) were more common in non-shockable cardiac arrests compared to shockable.^37^ A previous study from our group, which compared PEA to VF/VT cases, reported that individuals taking antipsychotic drugs had over double the risk of manifesting with PEA (OR 2.40).^11^ Similarly, we have also reported that subjects with a history of syncope or COPD had double the risk of manifesting with PEA compared to VF/VT.^31^ Our recent machine learning analysis among EMS-witnessed SCA cases identified anemia and kidney disease as determinants of PEA.^10^ These predictors are also components of PEA-Risk. In a South Korean nationwide cohort, anemia was independently associated with an increased risk of SCA, potentially related, at least in part, to QT interval prolongation.^38^ The correlation between anemia and SCA might be explained by an increase in arrhythmic risks, such as QTc prolongation.

Our findings have potential implications for revisiting the approach to PEA prevention and management. Lung and tissue hypoxia are consistent components of PEA pathophysiology. Conditions like COPD can lead to severe pulmonary compromise and hypoxia. Anemia results in hypoxia at the tissue level. Modification in resuscitation practices over time appear to have at least moderate beneficial effects on survival from PEA-SCA,^39^ also suggesting that specific subgroups of PEA cases may have better survival. Incremental severity of COPD is associated with increasing prevalence of non-shockable SCA.^40^ Preemptive management of hypoxia and anemia in individuals with high PEA-risk scores could reduce likelihood of PEA. Additionally, since antipsychotics and antidepressants increase the risk of PEA-SCA, these may have to be prescribed with care in individuals with high PEA-risk scores.

### Study Limitations

The identification of feasible numbers of out-of-hospital SCA cases manifesting with PEA mandates the use of a large, population-based design. Some inherent limitations of this type of study design should be considered while interpreting these findings. Due to the absence of established cardiac conditions or warning signs, a sizeable proportion of community residents may not have approached health care providers for evaluation before their unexpected SCA event. Others may have had variable types of clinical evaluations and treatments performed. Therefore, information regarding clinical profile and tests incorporated in the clinical scores was not uniformly available for all SCA cases. Secondly, we cannot be fully sure that PEA was the actual primary rhythm in all cases and not a result of spontaneous rhythm transition before the rhythm recording.

Thirdly, our AUC in the external validation was lower than our internal validation dataset 0.704 [0.665-0.743] vs 0.832 [0.800-0.865], and each point in the PEA-Risk score was associated with 13% lesser odds of predicting PEA as compared to the internal validation (OR 1.20 vs 1.33). This could be because our controls for the external validation were patients seeking health care who had a higher comorbidity burden than the internal validation controls, and median PEA-Risk scores were higher in the external validation controls than the training controls (6.7 vs 4.6).

## Conclusions

A risk score for prediction of sudden cardiac arrest manifesting with PEA was successfully constructed using widely available clinical and noninvasive markers. The algorithm was replicated in a geographically distinct population. This risk score performed equally well across subpopulations stratified by sex, age and coronary artery disease. These findings have implications for developing prevention and management strategies for PEA-SCA.

## Data Availability

Deidentified participant data will be made available after publication upon reasonable request to the corresponding author, following approval of a proposal and a signed data use agreement.

## Acknowledgments

We gratefully acknowledge the significant contributions of study participants from the Portland, OR metro area and Ventura County, CA.

## Supplemental Material

Tables S1–S2

## Non-standard Abbreviations and Acronyms

PEA-SCA: SCA presenting with documented PEA

---

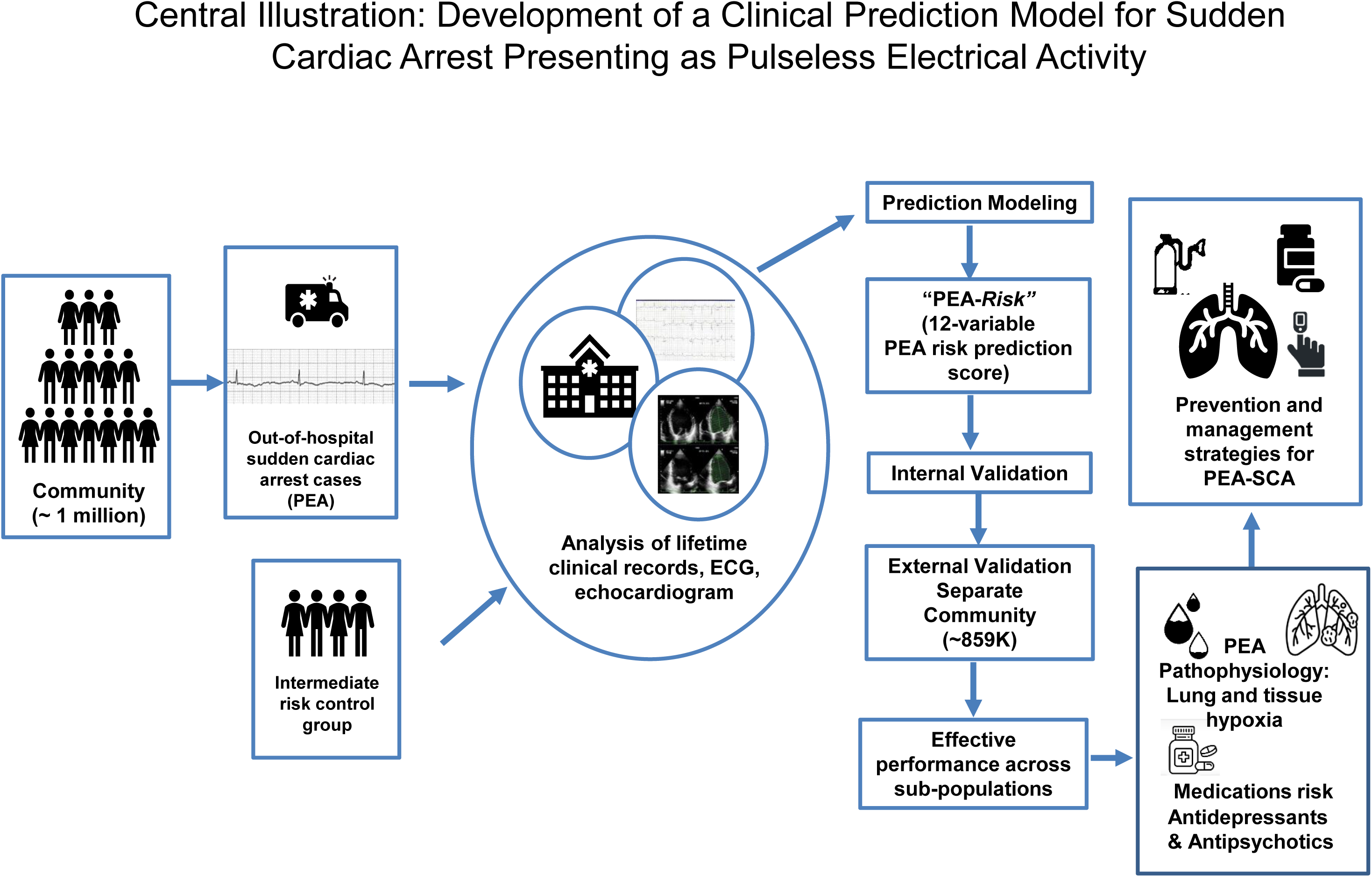

